# First Line Thrombectomy Devices in Intracranial Atherosclerotic Disease: An analysis of the RESCUE-ICAS registry

**DOI:** 10.1101/2025.08.06.25333179

**Authors:** Adam T Mierzwa, Ahmad abu Qdais, Imad Samman Tahhan, Shadi Yaghi, Violiza Inoa, Francesco Capasso, Michael Nahhas, Robert M. Starke, Isabel Fragata, Matthew Bender, Krisztina Moldovan, Ilko Maier, Jonathan A. Grossberg, Pascal Jabbour, Marios Psychogios, Edgar A Samaniego, Jan-Karl Burkhardt, Brian Jankowitz, Mohamad Abdalkader, David Altschul, Justin Mascitelli, Robert W. Regenhardt, Stacey Wolfe, Mohamad Ezzeldin, Kaustubh Limaye, Hosam Al Jehani, Hafeez Niazi, Nitin Goyal, Stavropoula Tjoumakaris, Ali Alawieh, Mohammed Almekhlafi, Eytan Raz, Syed Zaidi, Alejandro M. Spiotta, Kimberly Kicielinski, Jonathan Lena, Zachary Hubbard, Osama O. Zaidat, Colin P. Derdeyn, Ramesh Grandhi, Eyad Almallohi, Mohamad Anadani, Adam de Havenon, Thanh N. Nguyen, Ameer E. Hassan, Mohammad Jumaa, Sami Al Kasab

## Abstract

**Introduction:** Managing atherosclerotic large vessel occlusion is procedurally challenging. Prior literature pertaining to technical considerations remain heterogenous and further research is necessary to highlight important differences. As such, first-line thrombectomy technique remains an active area of debate with respect to rate of recanalization, need for rescue stenting, and hemorrhagic complications.

**Methods:** This is a pre-planned analysis of the prospective RESCUE-ICAS registry which included atherosclerotic large vessel occlusions treated with mechanical thrombectomy from 25 sites. Patients were excluded if they had missing data on first-line technique or primary outcomes. Patients were dichotomized into two cohorts based on whether their first-line thrombectomy technique was with aspiration alone or a stentriever (SR). Primary procedural outcome was first-pass effect while primary safety outcome was mortality at 90 days. Propensity score matching and inverse probability weighted analysis were performed with respect to primary and secondary outcomes.

**Results:** 419 were patients included in this analysis with 266 and 153 patients in the aspiration and stentriever cohorts, respectively. The cohort’s mean age was 68 (SD ±13) years, and the majority of patients were white (59%) and male (62%). There were no significant baseline demographic differences between cohorts; however, ICA occlusions were more common in the stentriever cohort (52% vs 31%), while MCA occlusions were more frequent in the aspiration cohort (35% vs 15%). In the un-adjusted model, first pass effect was higher in stentriever versus aspiration (35.3% vs 23.7%, p = 0.01) with equivalent mortality rates (31% vs 26%, p = 0.31). Distal embolization rates were higher in the aspiration cohort (9.8% vs 3.9%, p = 0.03), yet aspiration was associated with lower composited procedural complications (6% vs 11%, p = 0.01). Propensity score matching and weighted analysis demonstrated that differences in primary clinical efficacy and safety outcomes were insignificant between cohorts.

**Conclusion:** In patients with atherosclerotic large vessel occlusions, first line stentriever utilization was associated with higher first-pass effect rates, lower rates of distal embolization and shorter procedural length compared to aspiration. However, no clinical outcome difference was appreciated between the two groups and aspiration was associated with lower complication rates.

## Introduction

Mechanical thrombectomy (MT) for large vessel occlusion (LVO) ischemic stroke has been a mainstay therapy for nearly a decade.^1-5^ With the dramatic and rapid functional benefit of MT, stroke management underwent a revolutionary period in which clinicians and researchers have continued expanding opportunities to incorporate this treatment modality. Yet, other efforts are honed towards procedural optimization (e.g. door-to-puncture metrics, arteriotomy access, system composition, etc.). More recently, pros and cons of different thrombectomy techniques have been explored, and in general, the decision for first-line aspiration alone (ASP) versus stentriever utilization (STR) hinges on situational intricacies and interventionalists’ preferences.^6^

One such situation is in the setting of *in situ* atherosclerotic LVO which occurs in ∼6-30% of LVOs.^7^ Often, increased vessel tortuosity, arterial stenosis, recalcitrant underlying lesions, and high re-occlusion rates accompany atherosclerotic LVOs and accounts for the majority of thrombectomy failures.^8-10^ These procedural challenges ultimately lead to worsened functional outcomes and higher mortality rates.^10,11^ Therefore, utilizing devices which can appropriately navigate the vasculature, remove the thrombus, and limit vessel wall abrasion is paramount to a successful procedure. Prior literature demonstrates similar outcomes for different approaches. However, the ideal approach for *in situ* atherosclerotic LVOs remains unclear.^12-15^

In this study, we aim to evaluate the effect of MT technique on outcome among patients with atherosclerotic LVO ischemic strokes.

## Methods

This is an analysis of the RESCUE-ICAS registry whose rationale and study design has been previously described.^16^ In brief, RESCUE-ICAS is a prospectively designed registry which included non-disabled (Mrs ≤ 2) adults (age 18-80) with a confirmed atherosclerotic large vessel occlusion presenting within the 24-hour time window from last known well and treated with mechanical thrombectomy. Patient data were obtained from 25 thrombectomy capable or comprehensive stroke centers throughout the United States, Europe and Asia. Patients within the registry were excluded from this analysis if the primary endpoint or treatment method were not available. Individuals were dichotomized based on which first-line thrombectomy device was employed (ASP or STR). Combined ASP and STR techniques, colloquially referred to as “Solumbra,” were classified as STR.

A frequentist approach was utilized for this analysis. The primary procedural efficacy endpoint was first-pass effect (defined as a first pass resulting in TICI ≥ 2b). Secondary procedural efficacy endpoints included: first pass effect for both excellent recanalization (TICI ≥ 2c) and complete recanalization (TICI = 3), as well as final successful, excellent, and complete recanalization scores, procedural length, and total number of passes. Procedural safety endpoints included procedural complications and distal embolization. Procedural complications were defined as a composite of perforation, groin hematoma, stent-related, and re-occlusion. The clinical efficacy endpoint was defined as functional independence (mRS ≤ 2) at 90 days, with secondary clinical efficacy end points including favorable functional outcomes (mRS ≤ 3), median mRS, 24-hour NIHSS, and post-MT DWI lesion volume. The safety outcome was defined as mortality, while secondary safety outcomes included symptomatic ICH and any ICH.

Univariate analysis was performed as follows. Continuous variables were summarized as means and standard deviations or median and interquartile ranges, as appropriate. Categorical data was represented by counts and percentages. Parametric and nonparametric continuous data were analyzed with independent t-tests or Mann-Whitney U tests, respectively. Chi-squared analysis was utilized for comparing categorical percentages.

Unadjusted odds ratios were obtained for dichotomous primary and safety endpoints. Propensity score matching and weighted analysis was performed with replacement using nearest neighbor methods. Variables were included in the model if the composited absolute standardized mean difference was greater than 0.1. Subgroup analysis was additionally performed. The final model was additionally adjusted for occlusion site, intracranial angioplasty, and intracranial stent placement.

Statistical significance was defined as p value less than 0.05. Both IBM SPSS Statistics (IBM Corporation, New York, NY, Version 29.0.0) and R were utilized for this analysis. Packages included tidyverse (version 2.0.0), cobalt (4.5.5), and MatchIt (version 4.5.5).^17-19^

## Results

### Baseline Demographics

Four-hundred-fifty-one patients were included in this registry and 32 patients were excluded due to indeterminate first-line device or primary efficacy endpoint. Therefore, 419 patients were included in this analysis. (Figure 1.) Overall, patients were more often older (68 years-old SD 13) white (59%) men (62%). Comorbidities included hypertension (79%), dyslipidemia (49%), diabetes mellitus (39%), and atrial fibrillation (15%). Prior stroke was noted in 98 (23%) patients with 288 (69%) having mRS of 0. There were no significant baseline demographic differences between cohorts. (Table 1, Demographics)

**Table 1.**
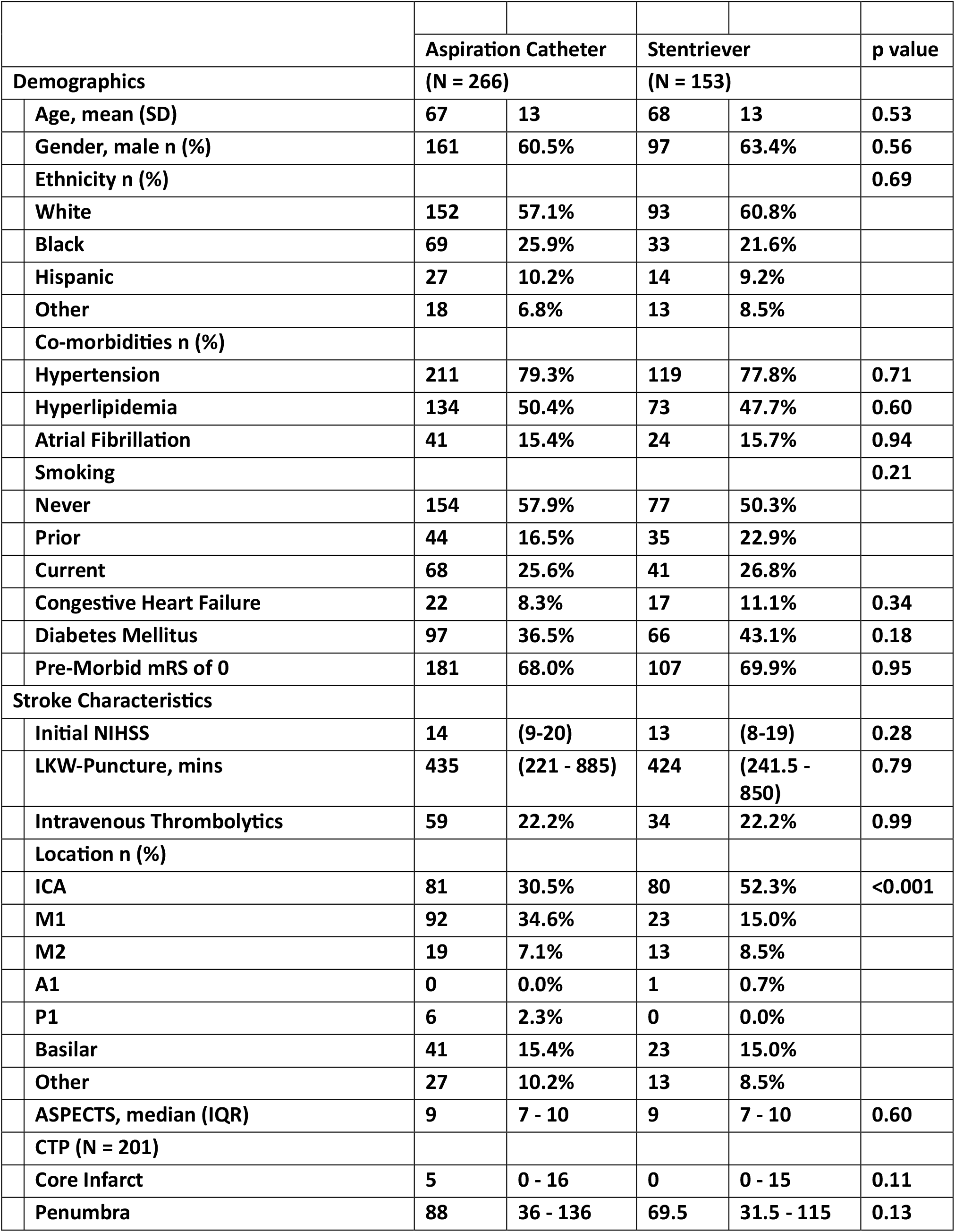
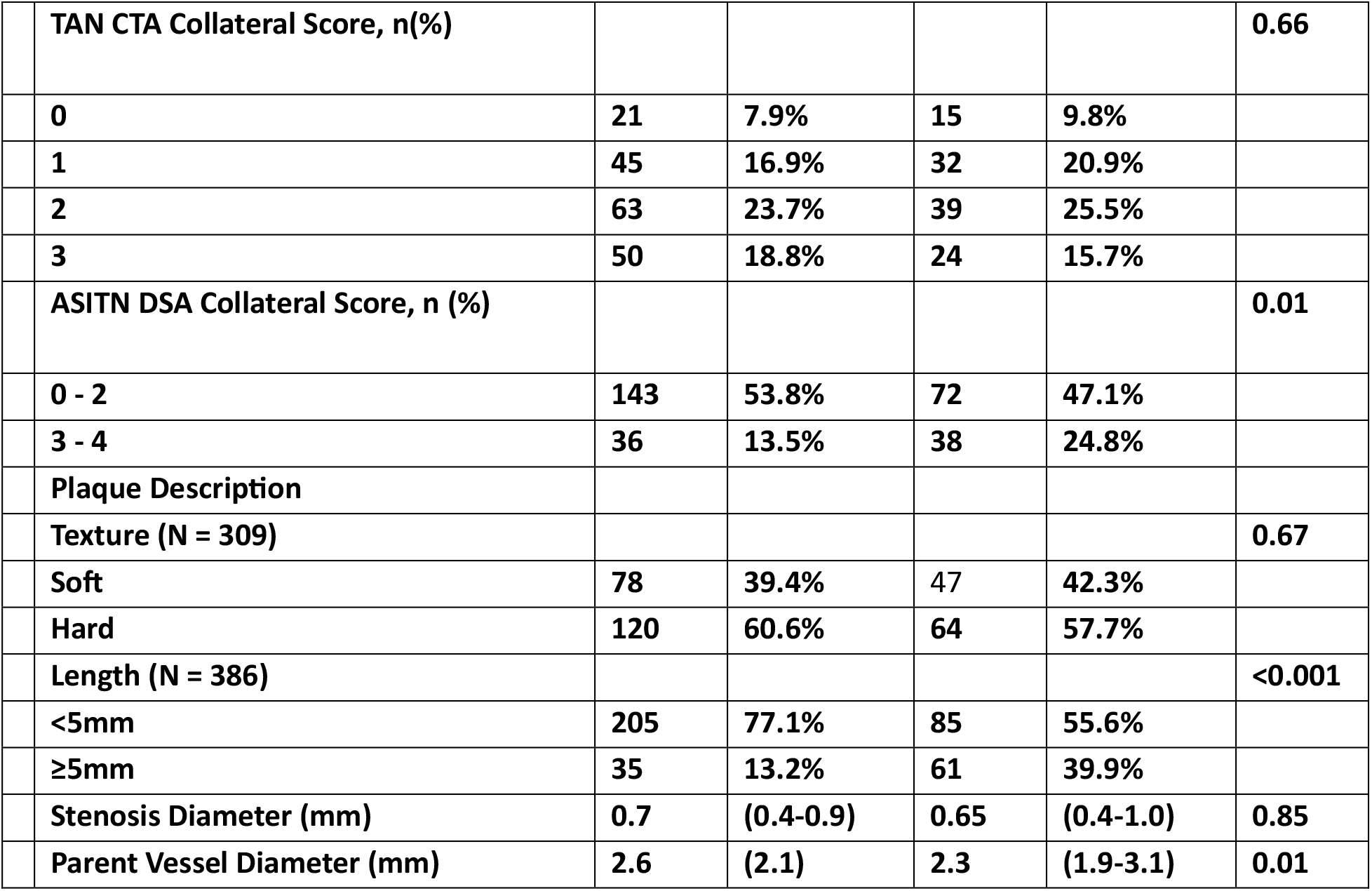
Patient Demographics and Stroke Characteristics.

| Table 1. Patient Demographics and Stroke Characteristics |  |  |  |  |  |  |
| --- | --- | --- | --- | --- | --- | --- |
|  |  | Aspiration Catheter |  | Stentriever |  | p value |
| Demographics |  | (N = 266) |  | (N = 153) |  |  |
|  | Age, mean (SD) | 67 | 13 | 68 | 13 | 0.53 |
|  | Gender, male n (%) | 161 | 60.5% | 97 | 63.4% | 0.56 |
|  | Ethnicity n (%) |  |  |  |  | 0.69 |
|  | White | 152 | 57.1% | 93 | 60.8% |  |
|  | Black | 69 | 25.9% | 33 | 21.6% |  |
|  | Hispanic | 27 | 10.2% | 14 | 9.2% |  |
|  | Other | 18 | 6.8% | 13 | 8.5% |  |
|  | Co-morbidities n (%) |  |  |  |  |  |
|  | Hypertension | 211 | 79.3% | 119 | 77.8% | 0.71 |
|  | Hyperlipidemia | 134 | 50.4% | 73 | 47.7% | 0.60 |
|  | Atrial Fibrillation | 41 | 15.4% | 24 | 15.7% | 0.94 |
|  | Smoking |  |  |  |  | 0.21 |
|  | Never | 154 | 57.9% | 77 | 50.3% |  |
|  | Prior | 44 | 16.5% | 35 | 22.9% |  |
|  | Current | 68 | 25.6% | 41 | 26.8% |  |
|  | Congestive Heart Failure | 22 | 8.3% | 17 | 11.1% | 0.34 |
|  | Diabetes Mellitus | 97 | 36.5% | 66 | 43.1% | 0.18 |
|  | Pre-Morbid mRS of 0 | 181 | 68.0% | 107 | 69.9% | 0.95 |
| Stroke Characteristics |  |  |  |  |  |  |
|  | Initial NIHSS | 14 | (9-20) | 13 | (8-19) | 0.28 |
|  | LKW-Puncture, mins | 435 | (221 - 885) | 424 | (241.5 - 850) | 0.79 |
|  | Intravenous Thrombolytics | 59 | 22.2% | 34 | 22.2% | 0.99 |
|  | Location n (%) |  |  |  |  |  |
|  | ICA | 81 | 30.5% | 80 | 52.3% | <0.001 |
|  | M1 | 92 | 34.6% | 23 | 15.0% |  |
|  | M2 | 19 | 7.1% | 13 | 8.5% |  |
|  | A1 | 0 | 0.0% | 1 | 0.7% |  |
|  | P1 | 6 | 2.3% | 0 | 0.0% |  |
|  | Basilar | 41 | 15.4% | 23 | 15.0% |  |
|  | Other | 27 | 10.2% | 13 | 8.5% |  |
|  | ASPECTS, median (IQR) | 9 | 7 - 10 | 9 | 7 - 10 | 0.60 |
|  | CTP (N = 201) |  |  |  |  |  |
|  | Core Infarct | 5 | 0 - 16 | 0 | 0 - 15 | 0.11 |
|  | Penumbra | 88 | 36 - 136 | 69.5 | 31.5 - 115 | 0.13 |

|  |  |  |  |  |  |
| --- | --- | --- | --- | --- | --- |
| TAN CTA Collateral Score, n(%) |  |  |  |  | 0.66 |
| 0 | 21 | 7.9% | 15 | 9.8% |  |
| 1 | 45 | 16.9% | 32 | 20.9% |  |
| 2 | 63 | 23.7% | 39 | 25.5% |  |
| 3 | 50 | 18.8% | 24 | 15.7% |  |
| ASITN DSA Collateral Score, n (%) |  |  |  |  | 0.01 |
| 0 - 2 | 143 | 53.8% | 72 | 47.1% |  |
| 3 - 4 | 36 | 13.5% | 38 | 24.8% |  |
| Plaque Description |  |  |  |  |  |
| Texture (N = 309) |  |  |  |  | 0.67 |
| Soft | 78 | 39.4% | 47 | 42.3% |  |
| Hard | 120 | 60.6% | 64 | 57.7% |  |
| Length (N = 386) |  |  |  |  | <0.001 |
| <5mm | 205 | 77.1% | 85 | 55.6% |  |
| ≥5mm | 35 | 13.2% | 61 | 39.9% |  |
| Stenosis Diameter (mm) | 0.7 | (0.4-0.9) | 0.65 | (0.4-1.0) | 0.85 |
| Parent Vessel Diameter (mm) | 2.6 | (2.1) | 2.3 | (1.9-3.1) | 0.01 |

**Table 2.** Outcomes.

| Table 2. Outcomes |  | Aspiration Catheter |  | Stentriever |  | p value |
| --- | --- | --- | --- | --- | --- | --- |
| Efficacy Outcomes |  | (N = 266) |  | (N = 153) |  |  |
|  | Procedural |  |  |  |  |  |
|  | FPE |  |  |  |  |  |
|  | TICI ≥ 2b | 63 | 23.6% | 54 | 35.3% | 0.01 |
|  | TICI ≥ 2C | 19 | 12.4% | 35 | 13.2% | 0.82 |
|  | TICI = 3 | 11 | 7.2% | 20 | 7.5% | 0.90 |
|  | Overall |  |  |  |  |  |
|  | TICI ≥ 2b | 224 | 84.2% | 128 | 83.7% | 0.88 |
|  | TICI ≥ 2c | 119 | 44.7% | 48 | 31.4% | 0.01 |
|  | TICI 3 | 61 | 22.9% | 25 | 16.3% | 0.11 |
|  | Procedural Length | 42 | (25-64) | 37 | (21-62) | 0.047 |
|  | Total # of Passes | 2 | (1-3) | 2 | (1-3) | 0.34 |
|  | Patient-Center |  |  |  |  |  |
|  | Functional Independence | 91 | 34.2% | 47 | 30.7% | 0.46 |
|  | Favorable Functional | 125 | 47.0% | 62 | 40.5% | 0.20 |
|  | median mRS | 4 | (2-6) | 4 | (2-6) | 0.39 |
|  | 24-Hour NIHSS | 13 | (4-20) | 10 | (5-20) | 0.06 |
|  | DWI-volume | 21 | (5-75) | 25 | (8-63) | 0.74 |

| Safety Outcomes |  |  |  |  |  |  |
| --- | --- | --- | --- | --- | --- | --- |
|  | Procedural |  |  |  |  | 0.01 |
|  | Perforation | 3 | 1.1% | 7 | 4.6% |  |
|  | Reocclusion | 6 | 2.3% | 3 | 2.0% |  |
|  | Dissection | 0 | 0.0% | 2 | 1.3% |  |
|  | Stent-related | 1 | 0.4% | 5 | 3.3% |  |
|  | Groin Hematoma | 5 | 1.9% | 0 | 0.0% |  |
|  | Distal Embolization | 26 | 9.8% | 6 | 3.9% | 0.03 |
|  | Patient-Center |  |  |  |  |  |
|  | Mortality | 82 | 30.8% | 40 | 26.1% | 0.31 |
|  | sICH | 18 | 6.8% | 14 | 9.2% | 0.38 |
|  | Total ICH | 71 | 26.7% | 29 | 19.0% | 0.21 |

**Figure 1.**
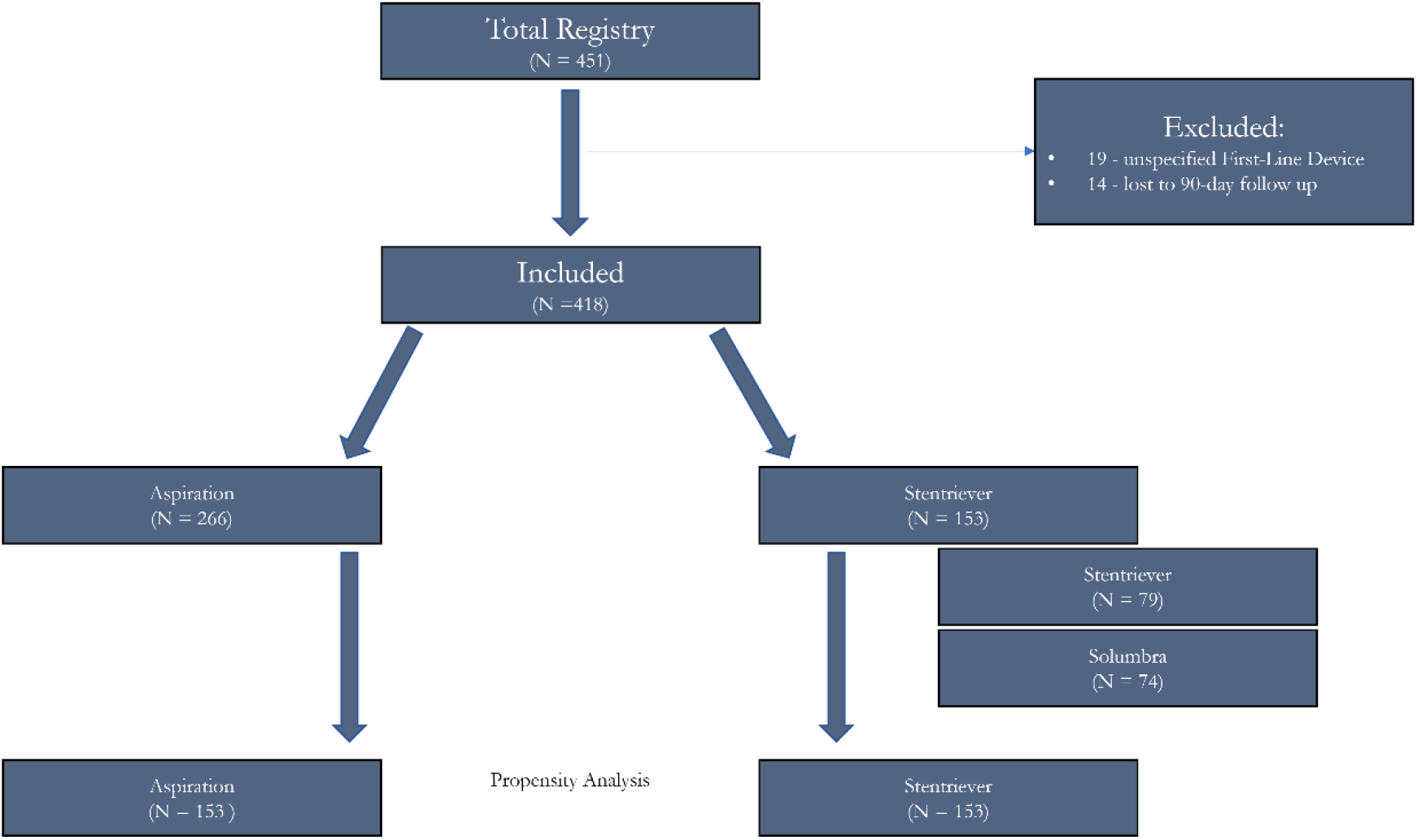
Patient Flow Chart

### Stroke Characteristics

Patients presented with a median NIHSS of 13 (IQR 9-19) with a median 425 (IQR 225 – 868) minutes from onset-to-puncture and 93 (22%) received intravenous thrombolysis. ASPECTS (9 IQR 7-10) as well as TAN collateral scores (61% ≥2) were favorable. The intracranial ICA was the most frequently occluded vessel (38%) followed by the MCA (27%). There was a soft underlying plaque in 40% of patients, with 75% of patients having an atherosclerotic lesion less than 5 mm in length. There were significant differences in the occluded segments (p<0.001) between cohorts with ASP having more frequent MCA occlusions (35% vs 15%) compared to STR cohort having more frequent ICA occlusions (52% vs 31%) (Table 1, Stroke Characteristics). Otherwise, no significant differences were noted.

### Efficacy Outcomes

ASP was less likely to obtain FPE compared to STR (23.6% vs 35.3%, OR 0.57, 95% CI 0.37 – 0.88; p = 0.01), while STR demonstrated shorter procedural length (37 mins vs 42 mins; p = 0.047). However, other procedural efficacy endpoints were equivalent between cohorts. (Table 3) Additionally, it was demonstrated that 76% of the ASP cohort required a second pass compared to 43% in the STR cohort (p <0.001). Furthermore, of the patients requiring a second thrombectomy attempt, it was more common for cases in the first-line ASP cohort to cross over to STR devices (47.4%) compared to those in the first-line STR cohort (15.7%) crossing over to an ASP device (p < 0.001). Yet, the STR cohort more often required balloon angioplasty (35.5% vs 24.4%; p = 0.01), but with similar rates of rescue stenting (44.4% vs 45.1%; p = 0.90) compared to the ASP cohort.

**Table 3.**
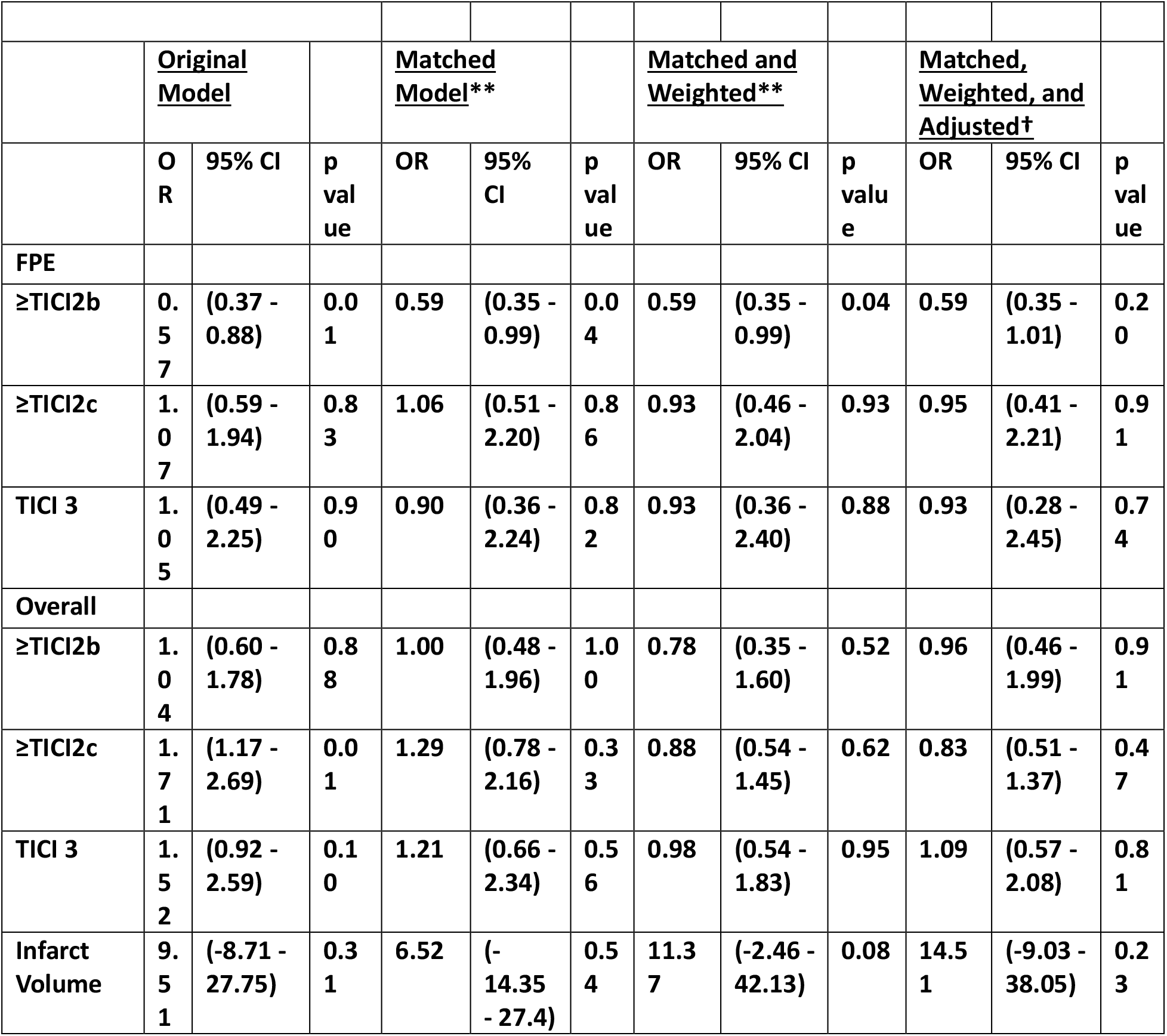
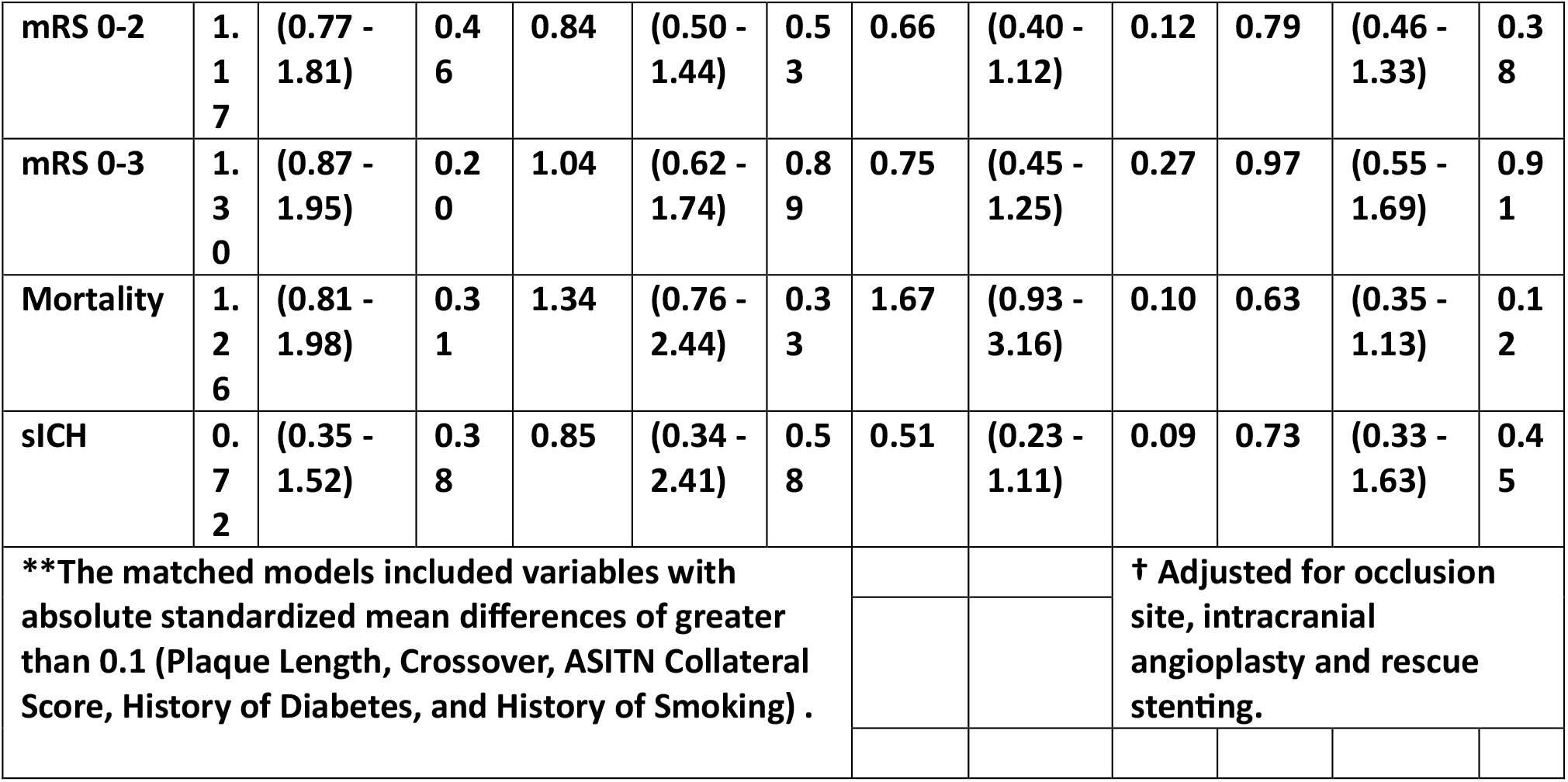
Odds Ratio Models.

There were 138 (32.9%) patients who achieved functional independence at 90-days, with 34.2% and 30.7% in the ASP and STR cohorts, respectively (OR 1.17, 95% CI 0.77 – 1.81; p = 0.46) (Figure 2). There was no significant difference in secondary efficacy outcomes including favorable functional (OR 1.30, 95% CI 0.87 – 1.95; p = 0.20), median mRS (4 vs 4; p = 0.39), 24-hour NIHSS (13 vs 10; p = 0.06), or post-MT DWI lesion volume (21 mL vs 25 mL; p = 0.74) between ASP and STR cohorts, respectively.

**Figure 2.**
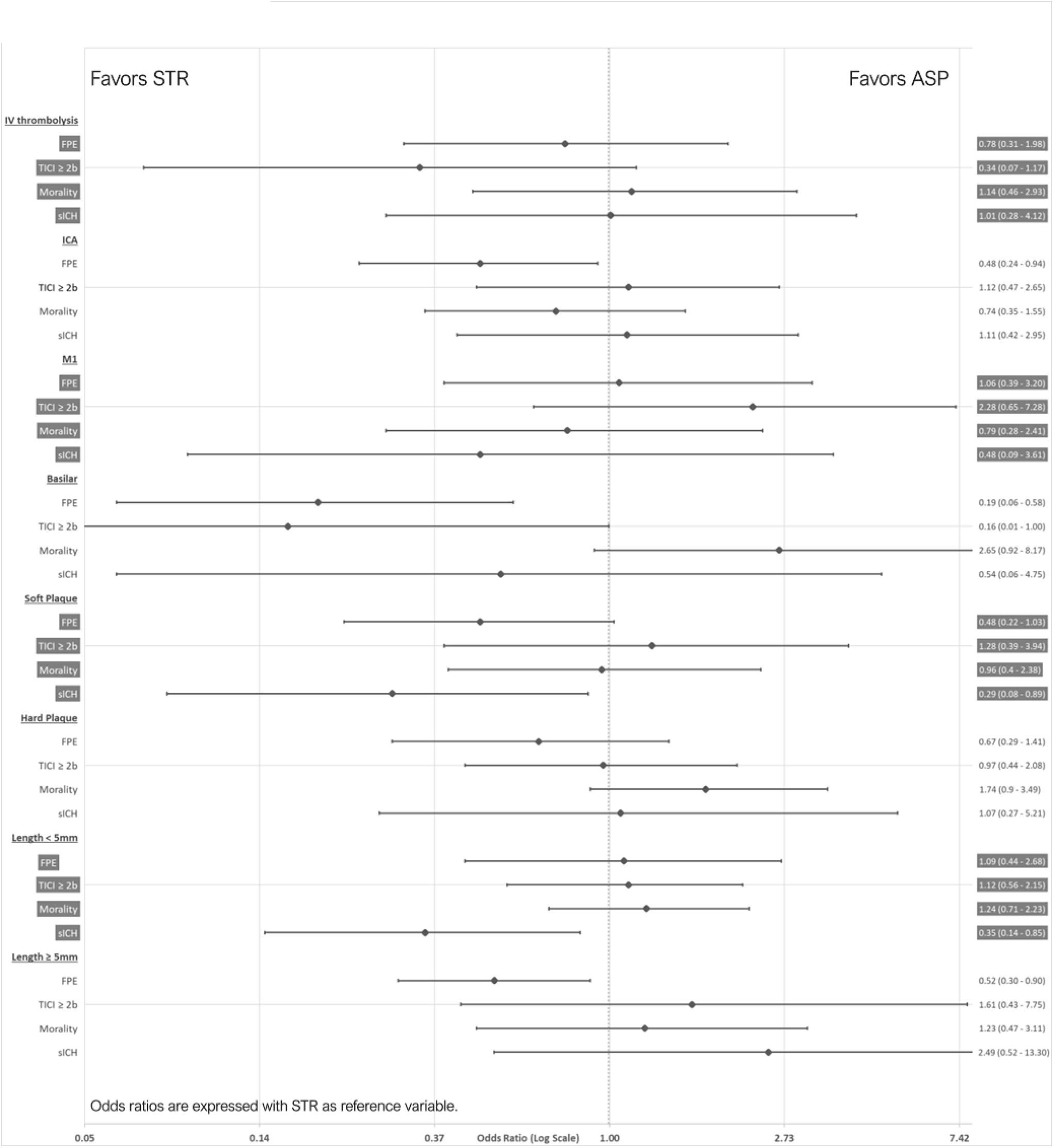
Subgroup Analaysis

### Safety Outcomes

Procedural complications were more common in the STR cohort, most notably perforation (4.6% vs 1.1%; p = 0.01), however, distal embolization was more common in the ASP cohort (9.8% vs 3.9%; p = 0.03).

Twenty-nine percent of patients met the other safety endpoint, with 82 (30.8%) and 40 (26.1%) in the ASP and STR cohorts, respectively (OR 1.26 95% CI 0.81 – 1.98; p = 0.31). There were similar rates of sICH between cohorts (6.8% vs 9.2%; p = 0.38) as well as any ICH (26.7% vs 19.0%; p = 0.21).

### Propensity Matching and Subgroups

ASITN collateral scores, history of diabetes, and history of smoking were included in the additional models. In the propensity matched cohort and weighted analysis, ASP and STR cohorts performed equally well in the primary or secondary efficacy or safety outcomes (Table 3, Figure 2). In the final multivariate model, functional and procedural efficacy outcomes remained similar. However, there was a trend between lower sICH rates with ASP compared to STR techniques (OR 0.73, 95% CI 0.33 – 1.63; p = 0.45).

## Discussion

This analysis of the RESCUE-ICAS prospective registry evaluated the influence of first-line thrombectomy technique on procedural efficacy, safety, and patient outcomes. We demonstrated that ASP and STR have similar functional independence rates in a direct model as well as in a propensity matched cohort and weighted analysis. This is similar to prior studies and further demonstrates that both ASP and STR techniques have a role for *in situ* atherosclerotic LVOs.^7,14^ The Effect of Endovascular Contact Aspiration vs Stent Retriever on Revascularization in Patients With Acute Ischemic Stroke and Large Vessel Occlusion (ASTER) trial directly compared ASP and STR techniques for LVO irrespective of associated ICAD, and demonstrated equivalent rates of favorable functional outcomes (45.3% vs 50.0%).^14^ This is further supported by the Aspiration thrombectomy versus stent retriever thrombectomy as first-line approach for large vessel occlusion (COMPASS) trial.^20^ However, this analysis and the COMPASS and ASTER trials have a key distinction in that this analysis only included patients with atherosclerotic LVOs. Furthermore, although there are similar functional outcomes in the aforementioned trial and this analysis, the underlying pathophysiology delineates procedural metrics.

First, previously described recanalization rates indicate ASP superiority or equivalency to STR techniques; however, this analysis demonstrated a lower first-pass effect (FPE) rate in the ASP cohort (23.7% vs 35.3%).^21,22^ The tortuous vasculature, stenosis, and fibrin-rich thrombi may overwhelm ASP maneuverability and impair adequate contact aspiration. Second, the ASP cohort demonstrated higher crossover rates (47% vs 16%) which is likely a pragmatic consequence from the first thrombectomy attempt. Third, although rescue stenting (44.4% vs 45.1%) was similar between groups, balloon angioplasty was more common (35.5% vs 24.4%) in the STR cohort. One interpretation is that the passage of the aspiration catheter through the plaque potentially acts as an angioplasty, diminishing the need of further balloon angioplasty later on. Also, STR techniques are more abrasive to the endothelial wall resulting in vessel architecture collapse requiring additional steps to restore flow. This concern is supported by an electron microscopy study in which post-thrombectomy stentrievers had vascular tissue embedded within the device.^23^ While a pilot study using high resolution MRI demonstrated that stentriever devices resulted in higher vessel wall enhancement as well as susceptibility signals in the surrounding stroke bed.^24^ Lastly, histopathological studies indicates aspiration catheters have a more favorable shear stress profile compared to stentrievers due to both adhesive and mechanical interactions.^25,26^ As a note of caution, despite not being appreciated in our series, is the risk of vessel dissection utilizing aspiration; while advancing forward the aspiration catheter during aspiration is a commonly used and safe technique for majority of thrombectomies, in setting of ICAD this technique may lead to the catheter advancing though the plaque creating a false lumen and a dissection.^27^

Mortality and symptomatic intracranial hemorrhage rates were similar between cohorts. This was further supported by the propensity matched and weighted analyses. Yet, there are several notable findings. On the secondary safety outcomes, the sICH rates demonstrated in this analysis were slightly higher (7.6%) compared to a previously published study level meta-analysis (5%).^15^ Additionally, this analysis determined significantly variable rates of procedural complications (5.6% vs 11.1%) and distal embolization (9.8% vs 3.9%) in the ASP and STR cohorts, respectively. These differences are likely related to the technical difficulties described previously as well as need for rescue therapy.

In a hypothesis generating subgroup analysis, it was surprising to demonstrate that first-line STR was associated with higher rates of symptomatic intracranial hemorrhages in short (OR 2.86 95% CI 1.17 – 7.14) and soft plaques (OR 1.67 95% CI 1.12 – 12.50) compared to ASP cohort. This is peculiar because this plaque morphology (Mori A) was considered favorable in a study performed by Kang et al.^28^ We consider that the lesions in this analysis and those described by Kang are fundamentally different. In the aforementioned study, patients were included if they experienced perfusion dependent ischemic strokes and were treated with an urgent intracranial stent (i.e. ≤14 days from qualifying event). These lesions in particular are likely stable plaques. However, the short and soft plaques experienced in this registry have evidently reached a volatile enough threshold to cause an LVO which indicates a highly inflammatory lesion and adventitial compromise. Therefore, thrombectomy strategy should be carefully selected when encountering aggressive plaque morphology.

This analysis has several limitations. First, radiographic data were not centrally adjudicated. This may improve generalizability, albeit at the cost of interrater bias. Second, many within the ASP cohort had low FPE and high crossover which limits direct cohort comparison of the overall approach while still directly comparing the first pass technique approach. Despite this, we advocate internal validity since our propensity matched and weighted analysis demonstrated similar primary and safety effect sizes which limits confounding. Third, although the data were prospectively obtained, patients were not randomized to treatment strategy, and further randomized clinical trials evaluating first-line device in atherosclerotic LVOs are required.

## Conclusion

Stentrievers have higher rates of first-pass effect and shorter procedural times for *in-situ* large vessel occlusions compared to aspiration catheters. However, first-line aspiration had more frequent complete recanalization rates and lower complications, including perforations. Despite these procedural differences, clinical outcomes were similar between cohorts.

## Data Availability

Data will be made available upon submission and approval of requests to corresponding author.

